# Effect of Vitamin D_3_ Supplementation vs Placebo on Hospital Length of Stay in Patients with Severe COVID-19: A Multicenter, Double-blind, Randomized Controlled Trial

**DOI:** 10.1101/2020.11.16.20232397

**Authors:** Igor H. Murai, Alan L. Fernandes, Lucas P. Sales, Ana J. Pinto, Karla F. Goessler, Camila S. C. Duran, Carla B. R. Silva, André S. Franco, Marina B. Macedo, Henrique H. H. Dalmolin, Janaina Baggio, Guilherme G. M. Balbi, Bruna Z. Reis, Leila Antonangelo, Valeria F. Caparbo, Bruno Gualano, Rosa M. R. Pereira

**Affiliations:** Rheumatology Division, Hospital das Clinicas HCFMUSP, Faculdade de Medicina da Universidade de Sao Paulo, Sao Paulo, Brazil; Applied Physiology & Nutrition Research Group; Faculdade de Medicina da Universidade de Sao Paulo, Sao Paulo, Brazi; Clinical Pathology Division, Hospital das Clinicas HCFMUSP, Faculdade de Medicina da Universidade de Sao Paulo, Sao Paulo, Brazil; Food Research Center, Universidade de Sao Paulo, Sao Paulo, Brazil

## Abstract

**Importance:** Patients with COVID-19 may exhibit 25-hydroxyvitamin D deficiency, but the beneficial effects of vitamin D_3_ supplementation in this disease remain to be proven by randomized controlled trials.

**Objective:** To investigate the efficacy and safety of vitamin D_3_ supplementation in patients with severe COVID-19.

**Design, Setting, and Participants:** This is a multicenter, double-blind, randomized, placebo-controlled trial conducted in two centers (a quaternary hospital and a field hospital) in Sao Paulo, Brazil. The trial included 240 hospitalized patients with severe COVID-19. The study was conducted from June 2, 2020 to October 7, 2020.

**Interventions:** Patients were randomly allocated (1:1 ratio) to receive either a single oral dose of 200,000 IU of vitamin D_3_ or placebo.

**Main Outcomes and Measures:** The primary outcome was hospital length of stay, defined as hospital discharge from the date of randomization or death. Secondary outcomes were mortality, admission to ICU, mechanical ventilation requirement, and serum levels of 25-hydroxyvitamin D, creatinine, calcium, C-reactive protein, and D-dimer.

**Results:** Of 240 randomized patients (mean age, 56 years; 56% men), 232 (96.7%) were included in the primary analysis. Log-rank test showed that hospital length of stay was comparable between the vitamin D_3_ supplementation and placebo groups (7.0 days [95% CI, 6.1 to 7.9] and 7.0 days [95% CI, 6.2 to 7.8 days]; hazard ratio, 1.12 [95% CI, 0.9 to 1.5]; *P* = .379; respectively). The rate of mortality (7.0% vs 5.1%; *P* = .590), admission to ICU (15.8% vs 21.2%; *P* = .314), and mechanical ventilation requirement (7.0% vs 14.4%; *P* = .090) did not significantly differ between groups. Vitamin D_3_ supplementation significantly increased serum 25-hydroxyvitamin D levels compared to placebo (difference, 24.0 ng/mL [95% CI, 21.0% to 26.9%]; *P =* .001). No adverse events were observed.

**Conclusions and Relevance:** Among hospitalized patients with severe COVID-19, vitamin D_3_ supplementation was safe and increased 25-hydroxyvitamin D levels, but did not reduce hospital length of stay or any other relevant outcomes vs placebo. This trial does not support the use of vitamin D_3_ supplementation as an adjuvant treatment of patients with COVID-19.

**Key points:** *Question:* Can vitamin D_3_ supplementation reduce hospital length of stay in hospitalized patients with severe COVID-19?

*Findings:* In this double-blind, randomized, placebo-controlled trial involving 240 hospitalized patients with severe COVID-19, a single dose of 200,000 IU of vitamin D_3_ supplementation was safe and effective in increasing 25-hydroxyvitamin D levels, but did not significantly reduce hospital length of stay (hazard ratio, 1.12) or any other clinically-relevant outcomes compared with placebo.

*Meaning:* Vitamin D_3_ supplementation does not confer therapeutic benefits among hospitalized patients with severe COVID-19.

## Introduction

A growing body of evidence has indicated that vitamin D may enhance the innate^1-3^ and adaptive immunity.^4, 5^ Since antigen-presenting cells have the ability to synthesize 1,25-dihydroxyvitamin D (the active form of vitamin D) from 25-hydroxyvitamin D, it has been postulated that vitamin D supplementation could improve the function of macrophages and dendritic cells, thereby ameliorating overall immune response.^6^ In fact, insufficient vitamin D status has been suggested as a potential risk factor for non-communicable^7^ and acute respiratory tract diseases,^8, 9^ including viral infections.^10^In this context, it has been recently conjectured that optimal levels of vitamin D could play important immunomodulatory and anti-inflammatory roles, thereby benefiting patients with COVID-19.^11, 12^ However, the putative benefits of supplementary vitamin D_3_ to patients with COVID-19 remain speculative and partially supported by limited data from observational studies and one small-scale, non-randomized clinical trial.^13-15^ To our knowledge, this is the first randomized, double-blind, placebo-controlled trial to investigate the safety and efficacy of vitamin D_3_ supplementation on hospital length of stay and other relevant clinical outcomes in hospitalized patients with severe COVID-19. Our main *a priori* hypothesis was that a single dose of 200,000 IU of vitamin D_3_ supplementation would increase 25-hydroxyvitamin D levels and shorten hospital length of stay among these patients.

## Methods

The study was approved by the Ethics Committee of Clinical Hospital of the School of Medicine of the University of Sao Paulo and by the Ethics Committee of Ibirapuera Field Hospital. All the procedures were conducted in accordance with the Declaration of Helsinki. The participants provided written informed consent before being enrolled in the study (Ethics Committee Approval Number 30959620.4.0000.0068). The trial protocol and statistical plan are included in Supplement 1. This manuscript was written according to the recommendations by the Consolidated Standards of Reporting Trials (CONSORT) guidelines (see Supplement 2).

### Participants

Hospitalized patients were recruited from Clinical Hospital of the School of Medicine of the University of Sao Paulo (a quaternary referral teaching hospital), and from Ibirapuera Field Hospital, both located in Sao Paulo, Brazil. Enrollment started on June 2, 2020, to August 27, 2020, with the final follow-up on October 7, 2020.

### Inclusion criteria

Inclusion criteria were: 1) adults aged 18 years or older; 2) diagnosis of COVID-19 by either polymerase chain reaction (PCR) for severe acute respiratory syndrome coronavirus 2 (SARS-CoV-2) from nasopharyngeal swabs or computed tomography scan findings (bilateral multifocal ground-glass opacities ≥ 50%) compatible with the disease; 3) diagnosis of flu syndrome with hospitalization criteria on hospital admission, presenting respiratory rate ≥ 24 breaths per minute, saturation < 93% on room air or risk factors for complications, such as heart disease, diabetes mellitus, systemic arterial hypertension, neoplasms, immunosuppression, pulmonary tuberculosis, and obesity, followed by COVID-19 confirmation before randomization.

### Exclusion criteria

Exclusion criteria were: 1) patient unable to read and sign the written informed consent; 2) patient already admitted under invasive mechanical ventilation; 3) previous vitamin D_3_ supplementation (> 1000 IU/day); 4) renal failure requiring dialysis or creatinine ≥ 2.0 mg/dL; 5) hypercalcemia defined by total calcium > 10.5 mg/dL; 6) pregnant or lactating women; and 7) patients with expected hospital discharge in less than 24 hours.

### Study design and treatment

This was a multicenter, double-blind, parallel-group, randomized placebo-controlled trial. Eligibility screening was performed between June 2, 2020 to July 21, 2020 at Clinical Hospital of the School of Medicine of the University of Sao Paulo (n = 122), and from July 22, 2020 to August 27, 2020 at Ibirapuera Field Hospital (n = 118). The final follow-up in both centers was on October 7, 2020. Eligible patients were assigned in a 1:1 ratio into either the vitamin D_3_ supplementation group or the placebo group. The randomization list was created using a computer-generated code, which was managed by a staff member who had no role in the study. We assessed patients’ clinical status, coexisting chronic diseases, demographic characteristics, self-reported body weight and height, and ethnicity on hospital admission. Outcomes were assessed at baseline and on hospital discharge or death records.

The vitamin D_3_ supplementation group received an oral, single dose of 200,000 IU of vitamin D_3_ dissolved in a 10 mL of peanut oil solution on the same day of randomization. The selected dose is within the recommended range for effectively promoting vitamin D sufficiency.^16^ Patients in the placebo group received 10 mL of peanut oil solution. The vitamin D_3_ and placebo solutions were identical in color, taste, smell, consistency, and container. Both were prepared by the pharmacy unit of Clinical Hospital and labeled by a staff member who did not participate in the study. Allocation blindness was kept until the final statistical analysis.

### Outcome measures

The primary outcome was hospital length of stay, defined as the total number of days that patients remained hospitalized from the date of study admission until the date of hospital discharge or death. The criteria used for patient discharge were: 1) no need for supplemental oxygen in the last 48 hours; 2) no fever in the last 72 hours; and 3) oxygen saturation > 93% in room air without respiratory distress.

The secondary outcomes were: 1) mortality; 2) number of patients admitted to the intensive care unit (ICU); 3) number of patients who needed mechanical ventilation and duration of mechanical ventilation; and 4) serum levels of 25-hydroxyvitamin D (assessed by a chemiluminescent immunoassay), calcium (assessed by a NM-BAPTA method), creatinine (assessed by a colorimetric assay based on kinetic Jaffe’s reaction), and C-reactive protein and D-dimer (both assessed by an immunoturbidimetric assay). The biochemical analyses were carried out in an accredited laboratory from Clinical Hospital.

### Statistical Analysis

Considering the lack of data available for sample size determination based on the primary outcome (i.e., hospital length of stay after vitamin D_3_ supplementation in patients with severe COVID-19), the number of participants was chosen on the basis of feasibility, such as resources, capacity of research staff and facility, and available patients, in line with current recommendations.^17, 18^ Subsequently, we calculated sample size assuming a 50% between-group difference in hospital length of stay (considering 7 days as a median time of stay, with an expected variability of 9 days). By considering a power of 80% and a 2-sided significance level of 5% (α = .05), the total sample was estimated to be 208 patients (104 in each arm). Considering possible dropouts, and to increase the power for secondary outcomes, we opted by increasing the sample size by approximately 15%.

All analyses were carried out following the intention-to-treat principle for all randomized patients, with no imputation for any missing data. Proportions were compared between groups using χ^2^ test and Fisher’s exact test. Student’s t-tests were used for comparing continuous variables at baseline. The log-rank test was used to compare the Kaplan-Meier estimate curves the number of days for hospital length of stay, the primary outcome. Cox regression models for hospital length of stay, admission to ICU and mechanical ventilation requirement were adjusted by potential confounders that were not fully balanced by randomization (*P* < .2) to estimate hazard ratios (HR), with corresponding 2-sided 95% CI. Generalized estimating equations (GEE) for repeated measures were used for testing possible differences in laboratory parameters, assuming group and time as fixed factors, with marginal distribution, and a first-order autoregressive correlation matrix to test the main and interaction effects. *Post-hoc* tests with Bonferroni’s adjustment were performed for multiple comparisons. The aforementioned statistical procedures were also carried out in p*ost-hoc* sensitivity analyses involving patients exhibiting 25-hydroxyvitamin D deficiency (i.e., < 20 ng/mL). Statistical analyses were performed with IBM-SPSS software, version 20.0. Significance level was set at α = .05.

## Results

### Patients

Of 1208 patients assessed for eligibility, 240 were eligible and randomly assigned to either the vitamin D_3_ group or the placebo group. Patients were non-eligible due to the following reasons: 284 were at ICU, 263 had hospital discharge within 24 hours, 217 did not have COVID-19 confirmation, 95 had renal dysfunction, 37 had dementia or severe mental confusion hampering their ability to provide the inform consent for participation, 30 were pregnant or lactating women, 14 had hypercalcemia due to metastatic neoplasm, 11 were receiving vitamin D_3_ (≥ 1000 IU/day), 9 were younger than 18 years, 6 were illiterate and, therefore, unable to read and sign the informed consent, and 2 died before randomization.

Of the 120 patients who were randomized to the vitamin D_3_ group, 3 did not receive intervention (1 withdrew the consent, 1 vomited immediately after ingesting the supplement, and 1 was admitted to the ICU before taking vitamin D_3_) and 3 were lost to follow-up. Of the 120 patients who were randomized to the placebo group, 2 withdrew the consent. Thus, of the 240 patients randomized, 232 (96.7%) completed the follow-up (**Figure 1**).

**Figure 1.**
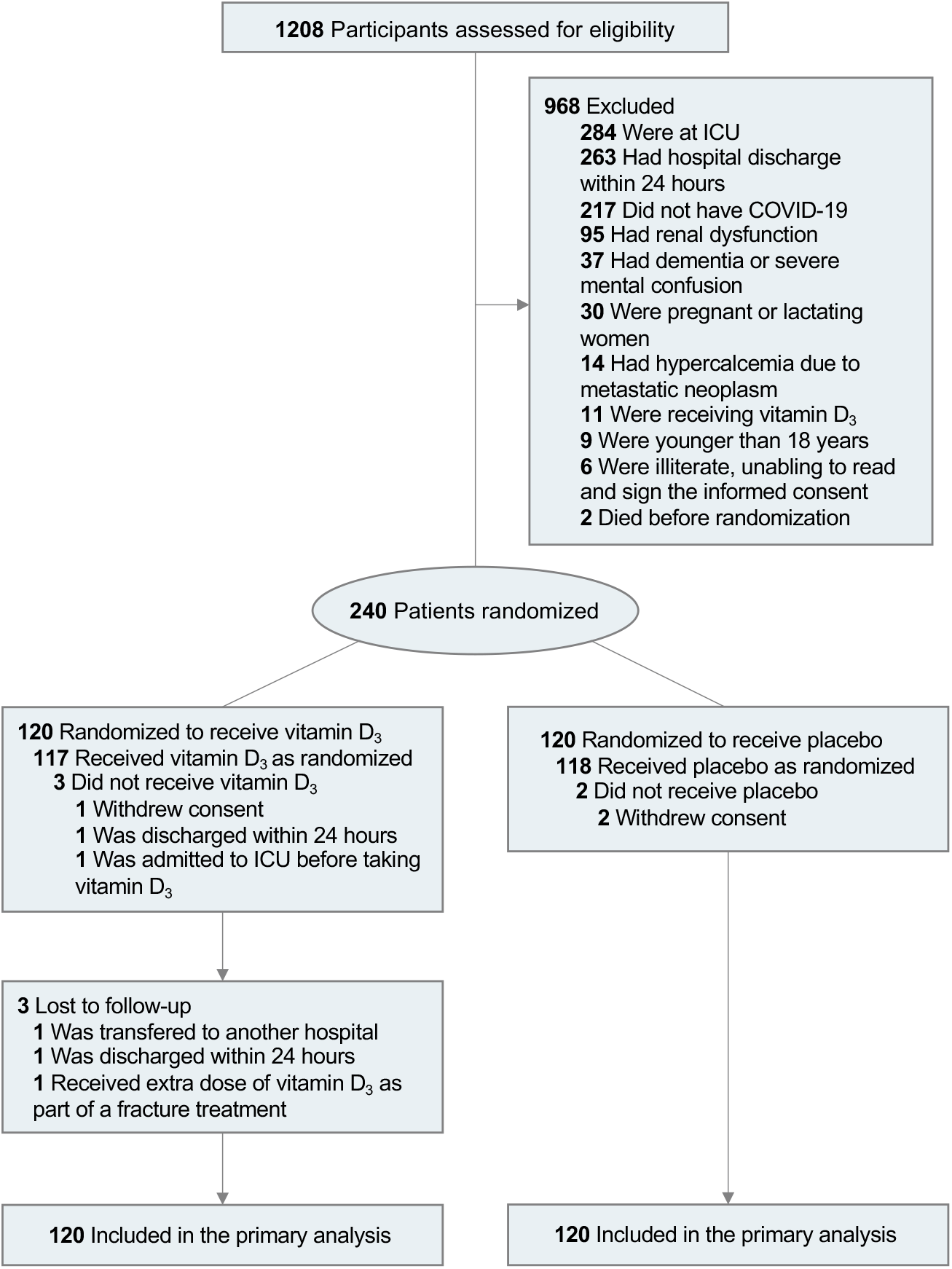
Flow of patients.

Overall, patients’ age was 56.3 years (SD, 14.6), BMI was 31.6 kg/m^2^ (SD, 7.1), 56.3% were men, 55% were white, 52.5% had hypertension, 35% had diabetes, 13.3% had cardiovascular diseases, and 6.3% had asthma. The mean time between the onset of symptoms and randomization was 10.2 days (SD, 4.3); 89.6% required supplemental oxygen at baseline (183 were on oxygen therapy and 32 were on non-invasive ventilation), and 59.6% had computed tomography scan findings suggestive of COVID-19. Demographic and clinical characteristics did not significantly differ between groups, except for sore throat, which was more prevalent in the vitamin D_3_ group vs placebo (38.3% vs 24.2%, *P* = .026), and PTH, which was higher in the vitamin D_3_ group vs placebo (50.1 vs 42.6 pg/mL, *P* = .025) (**Table 1**).

**Table 1.** Baseline demographic and clinical characteristics.

|  | Vitamin D<sub>3</sub><br>(n = 120) | Placebo<br>(n = 120) | *P*<br>value |
| --- | --- | --- | --- |
| Age, mean (SD), y | 56.8 (14.2) | 55.8 (15.0) | .584 |
| Sex, No. (%) |  |  |  |
| Male | 70 (58.3) | 65 (54.2) | .515 |
| Female | 50 (41.7) | 55 (45.8) |  |
| Race, No. (%) |  |  |  |
| White | 62 (51.7) | 70 (58.3) | .399 |
| Brown | 37 (30.8) | 36 (30.0) |  |
| Black | 20 (16.7) | 14 (11.7) |  |
| Asian | 1 (0.8) | 0 (0.0) |  |
| Days since symptoms onset, mean (SD) | 10.3 (4.7) [n=116] | 10.2 (3.8) [n=119] | .787 |
| Body mass index, mean (SD), kg/m<sup>2</sup> | 31.9 (6.5) [n=109] | 31.3 (7.6) [n=110] | .548 |
| Underweight, No./total (%) | 0 (0) | 2 (1.8) | .067 |
| Normal, No./total (%) | 9/109 (8.3) | 19/110 (17.3) |  |
| Overweight, No./total (%) | 37/109 (33.9) | 31/110 (28.2) |  |
| Obesity, No./total (%) | 63/109 (57.8) | 58/110 (52.7) |  |
| Acute COVID-19 symptoms, No. (%) |  |  |  |
| Fever | 86 (71.7) | 81 (67.5) | .575 |
| Cough | 103 (85.8) | 99 (82.5) | .596 |
| Fatigue | 98 (81.7) | 100 (83.3) | .865 |
| Joint pain | 46 (38.3) | 35 (29.2) | .172 |
| Myalgia | 69 (57.5) | 71 (59.2) | .896 |
| Nasal congestion | 39 (32.5) | 43 (35.8) | .683 |
| Runny nose | 44 (36.7) | 44 (36.7) | >.999 |
| Sore throat | 46 (38.3) | 29 (24.2) | .026 |
| Diarrhea | 41 (34.2) | 46 (38.3) | .591 |
| Coexisting diseases, No. (%) |  |  |  |
| Hypertension | 68 (56.7) | 58 (48.3) | .196 |
| Cardiovascular disease | 16 (13.3) | 16 (13.3) | >.999 |
| Diabetes | 49 (40.8) | 35 (29.2) | .058 |
| Chronic obstructive pulmonary disease | 7 (5.8) | 5 (4.2) | .554 |
| Asthma | 8 (6.7) | 7 (5.8) | .790 |
| Chronic kidney disease | 2 (1.7) | 0 (0.0) | .489 |
| Rheumatic disease | 13 (10.8) | 10 (8.3) | .511 |
| Concomitant medications, No. (%) |  |  |  |
| Antibiotic | 102 (85.0) | 105 (87.5) | .708 |
| Anticoagulant | 110 (91.7) | 103 (85.8) | .220 |
| Analgesic | 45 (37.5) | 52 (43.7) | .430 |
| Corticosteroids | 77 (64.2) | 73 (60.8) | .689 |
| Antihypertensive | 67 (55.8) | 56 (46.7) | .196 |
| Hypoglycemic | 26 (21.7) | 24 (20.0) | .874 |
| Hypolipidemic | 15 (12.5) | 18 (15.0) | .708 |
| Antiemetic | 45 (37.5) | 55 (45.8) | .239 |
| Antiviral | 4 (3.3) | 4 (3.3) | >.999 |
| Proton pump inhibitor | 47 (39.2) | 49 (40.8) | .895 |
| Thyroid | 10 (8.3) | 10 (8.3) | >.999 |
| Oxygen supplementation, No. (%) |  |  |  |
| No oxygen therapy | 16 (13.3) | 9 (7.5) |  |
| Oxygen therapy | 86 (71.7) | 97 (80.8) | .210 |
| Non-invasive ventilation | 18 (15.0) | 14 (11.7) |  |
| Computed tomography findings, No. (%) |  |  |  |
| Ground-glass opacities < 50% | 61 (50.8) | 66 (55.0) |  |
| Ground-glass opacities ≥ 50% | 47 (39.2) | 39 (32.5) | .543 |
| Not available | 12 (10.0) | 15 (12.5) |  |
| Laboratory variables |  |  |  |
| Haemoglobin, mean (SD), g/L | 13.1 (2.1) | 12.8 (2.1) | .298 |
| Neutrophils count, mean (SD),<br>x10<sup>3</sup>/mm<sup>3</sup> | 6.7 (4.0) [n = 119] | 7.2 (3.6) [n = 120] | .281 |
| Lymphocyte count, mean (SD),<br>x10<sup>3</sup>/mm<sup>3</sup> | 1.2 (0.5) [n = 119] | 1.1 (0.8) [n = 120] | .849 |
| Platelet count, mean (SD), x10<sup>3</sup>/mm<sup>3</sup> | 305.4 (116.4) | 286.7 (128.5) | .239 |
| Erythrocyte sedimentation rate, mean<br>(SD), mm | 58.4 (40.6) [n = 117] | 60.9 (36.7) [n = 119] | .627 |
| C-reactive protein, mean (SD), mg/L | 79.6 (75.2) [n = 119] | 90.4 (80.4) [n = 120] | .286 |
| D-dimer, mean (SD), ng/mL | 2091 (5001.3)<br>[n = 119] | 1720.7 (3630.1)<br>[n = 119] | .514 |
| Albumin, mean (SD), g/L | 3.1 (0.5) [n = 110] | 3.0 (0.4) [n = 100] | .454 |
| Gamma globulins, mean (SD), g/L | 1.1 (0.4) [n = 110] | 1.1 (0.3) [n = 100] | .337 |
| Creatinine, mean (SD), mg/dL | 0.9 (0.3) | 0.8 (0.2) | .149 |
| Urea, mean (SD), mg/dL | 40.1 (20.1) [n = 120] | 37.8 (14.5) [n = 119] | .309 |
| Phosphorus, mean (SD), mg/dL | 3.0 (0.6) [n = 117] | 3.0 (0.8) [n = 116] | .790 |
| 25-hydroxyvitamin D, mean (SD), ng/mL | 21.0 (10.2) [n = 118] | 20.6 (8.1) [n = 118] | .747 |
| Parathyroid hormone, mean (SD), pg/mL | 50.1 (27.3) [n = 113] | 42.6 (21.5) [n = 110] | .025 |
| Calcium, total, mean (SD), mg/dL | 8.7 (0.5) [n = 118] | 8.7 (0.5) [n = 119] | .811 |
| Total cholesterol, mean (SD), mg/dL | 164.3 (43.3) [n = 116] | 164.5 (47.2) [n = 115] | .975 |
| LDL-cholesterol, mean (SD), mg/dL | 101.3 (34.6) [n = 116] | 99.3 (39.1) [n = 114] | .681 |
| HDL-cholesterol, mean (SD), mg/dL | 34.6 (11.4) [n = 116] | 34.5 (11.0) [n = 114] | .916 |
| Triglycerides, mean (SD), mg/dL | 178.6 (75.5) [n=116] | 192.5 (93.6) [n = 114] | .218 |

For continuous variables, groups were compared using independent t-test.
For categorical variables, groups were compared using $\chi^2$ test or Fisher's exact test, as appropriate.

### Primary Outcome

Hospital length of stay **(Figure 2)** was comparable between the vitamin D_3_ group and the placebo group (7.0 days [95% CI, 6.1 to 7.9] and 7.0 days [95% CI, 6.2 to 7.8 days], HR, 1.12, [95% CI, 0.9 to 1.5]; *P* = .379; respectively). The Cox regression model did not show any significant associations between this outcome and potential confounders.

**Figure 2.**
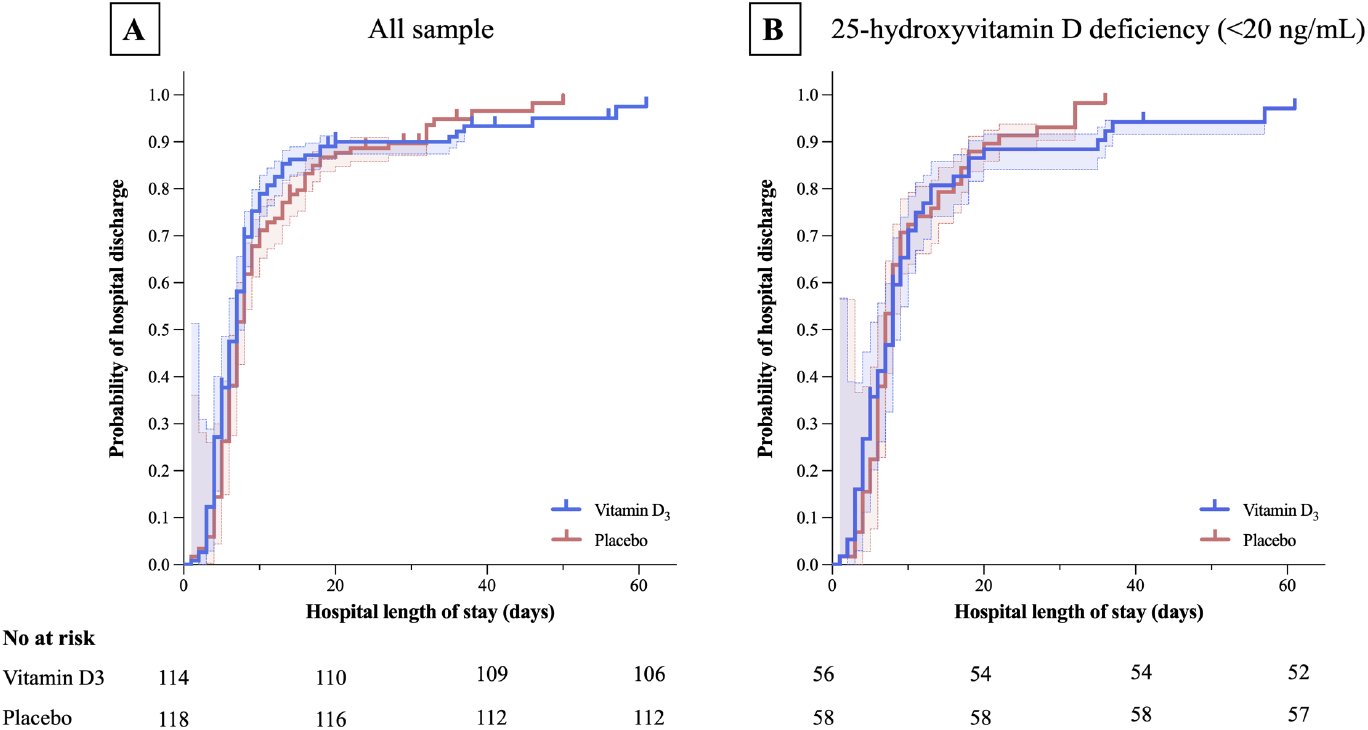
Kaplan-Meier curves for hospital length of stay. Vertical bars present single censored events. The shaded areas represent the 95% confidence intervals. The adjusted hazard ratio for total number of days that patients remained hospitalized from the date of study admission until the date of hospital discharge or death was 1.12 (95% CI, 0.9 to 1.5; *P* = .379) for the vitamin D_3_ group (7.0 days [95% CI, 6.1 to 7.9]) vs the placebo group (7.0 days [95% CI, 6.2 to 7.8 days]).

### Secondary Outcomes

There were no significant differences between the vitamin D_3_ group and the placebo group for mortality (7.0% vs 5.1%; *P* = .590), admission to ICU (15.8% vs 21.2%; *P* = .314) and mechanical ventilation requirement (7.0% vs 14.4%; *P* = .090) (**Figure 3**). Duration of mechanical ventilation was also comparable between the vitamin D_3_ group (18.1 days [95% CI, 3.5 to 32.7]) and the placebo group (11.4 days [95% CI, 7.1 to 15.6]; *P* = .549, respectively).

The Cox regression model did not show significant associations between secondary outcomes and potential confounders.

**Figure 3.**
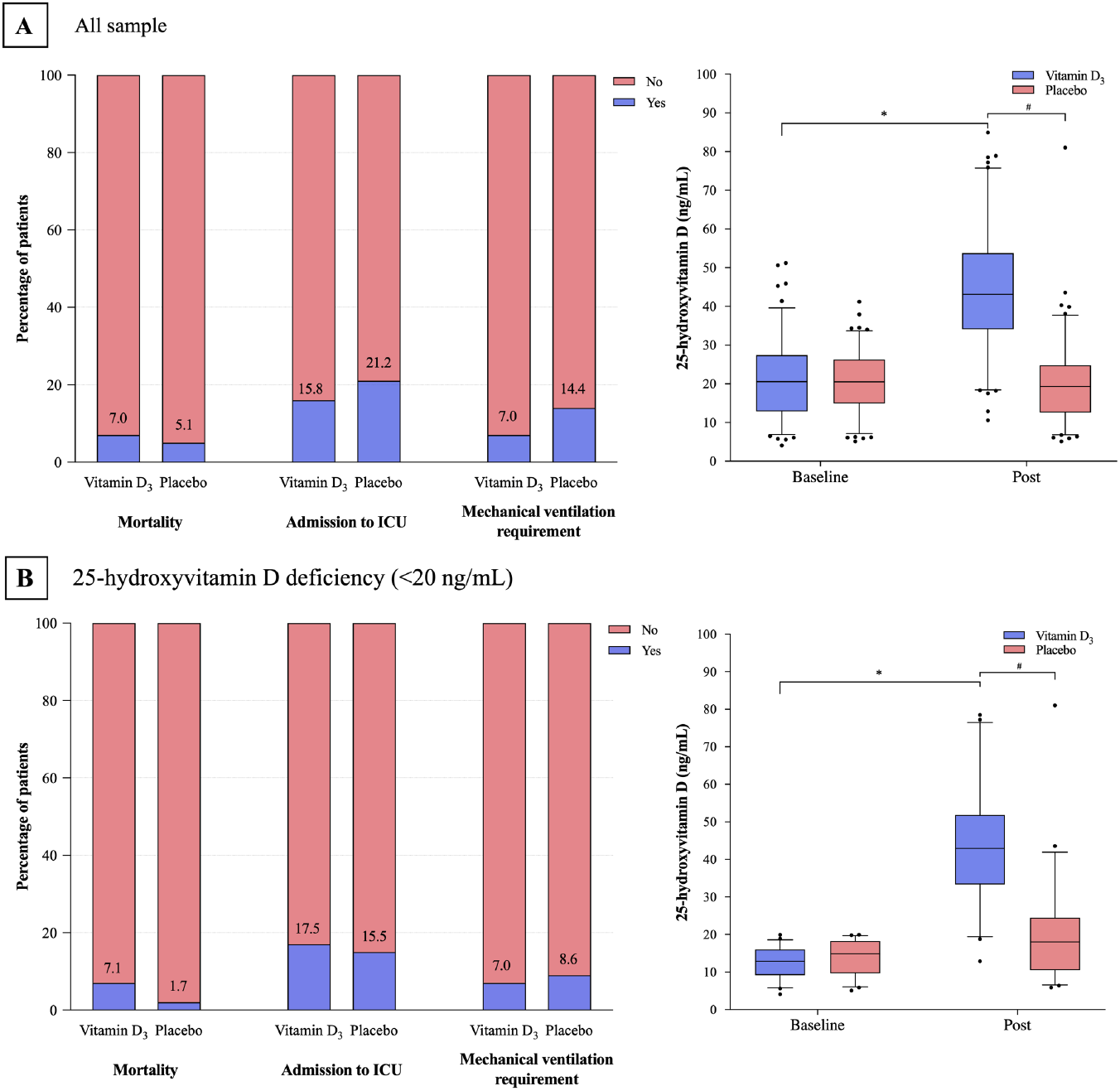
Serum 25-hydroxyvitamin D levels, mortality, admission to intensive care unit (ICU), and mechanical ventilation requirement. The panels show the comparisons between the vitamin D_3_ and placebo group for all patients (n = 240) (Panels A and B) and for those with 25-hydroxyvitamin D deficiency (< 20 ng/mL) (n = 116) (Panels C and D). For all patients, no significant differences were found between the vitamin D_3_ group and the placebo group for mortality (7.0% vs 5.1%; *P* = .590), admission to ICU (15.8% vs 21.2%; *P* = .314), and need of mechanical ventilation (7.0% vs 14.4%; *P* = .090). Vitamin D_3_ supplementation significantly increased 25-hydroxyvitamin D levels vs placebo (difference, 24.0 ng/mL [95% CI, 21.1-26.9]; *P <* .001). For patients with 25-hydroxyvitamin D deficiency, there were no significant differences between the vitamin D_3_ group and the placebo group for mortality (7.0% vs 1.7%; *P =* .206), admission to ICU (17.5% vs 15.5%; *P* = .806), and mechanical ventilation requirement (7.0% vs 8.6%; *P* > .999). Vitamin D_3_ supplementation significantly increased 25-hydroxyvitamin D levels vs placebo (difference, 22.7 ng/mL [95% CI, 19.3- 26.1]; *P* < .001). Box plots depict median and interquartile range. Outliers (i.e., defined as a value < 5 or > 95 percentiles) were represented by filled circles. * means *P* < .05 between Baseline and Post; ^#^ means *P* < .05 between groups at Post.

Vitamin D_3_ supplementation significantly increased 25-hydroxyvitamin D levels vs placebo (difference, 24.0 ng/mL [95% CI, 21.1-26.9]; *P <* .001) (**Figure 3**). Following the intervention, 86.7% of the patients in the vitamin D_3_ group showed 25-hydroxyvitamin D levels above 30 ng/mL (vs 10.9% in the placebo group), and only 6.7% of the patients in the vitamin D_3_ group exhibited 25-hydroxyvitamin D deficiency (vs 51.5% in the placebo group).

### *Post-hoc* Sensitivity Analyses

In a sensitivity analysis involving patients with 25-hydroxyvitamin D deficiency at baseline (n = 116) **(Supplementary Table 1)**, vitamin D_3_ supplementation significantly increased 25-hydroxyvitamin D levels vs placebo (difference, 22.7 ng/mL [95% CI, to 26.1]; *P* < .001) (**Figure 3**). Among the patients with 25-hydroxyvitamin D deficiency, no between-group differences were observed in length of hospital stay (**Figure 2**). In addition, there were no significant differences between the vitamin D_3_ group and the placebo group for mortality (7.0% vs 1.7%; *P* = .206), admission to ICU (17.5% vs 15.5%; *P* = .806), and mechanical ventilation requirement (7.0% vs 8.6%; *P* > .999) (**Figure 3**). Duration of mechanical ventilation did not differ between the vitamin D_3_ group (15.0 days [95% CI, -12.0 to 42.0]) and the placebo group (12.6 days [95% CI, -7.6 to 26.0]; *P* = .730).

### Safety and Adverse Events

There were no changes in any health-related laboratory markers following the intervention (**Table 2**). Vitamin D_3_ supplementation was well tolerated and no severe adverse events were reported throughout the trial, with the exception of one patient who vomited following vitamin D_3_ administration.

**Table 2.**
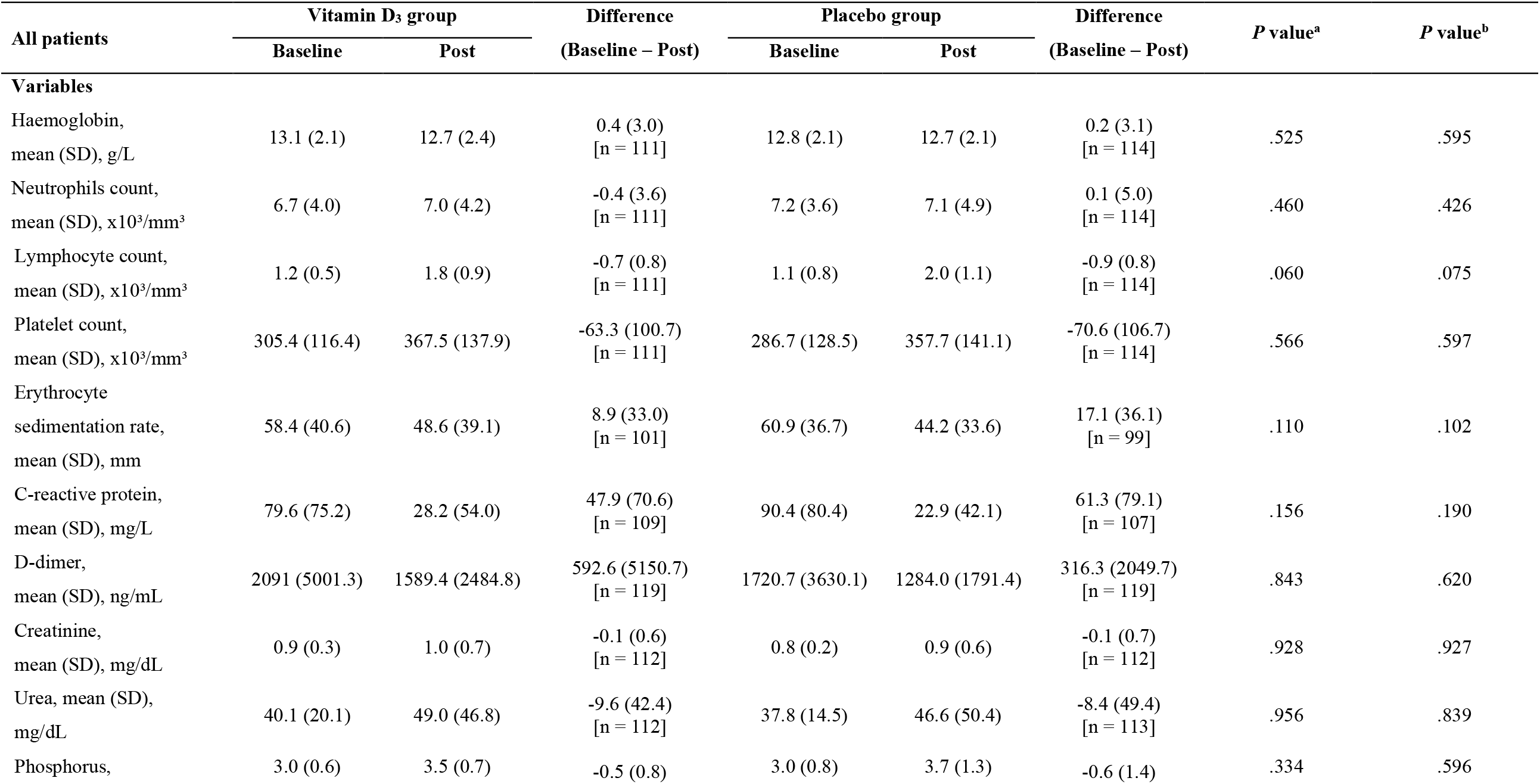
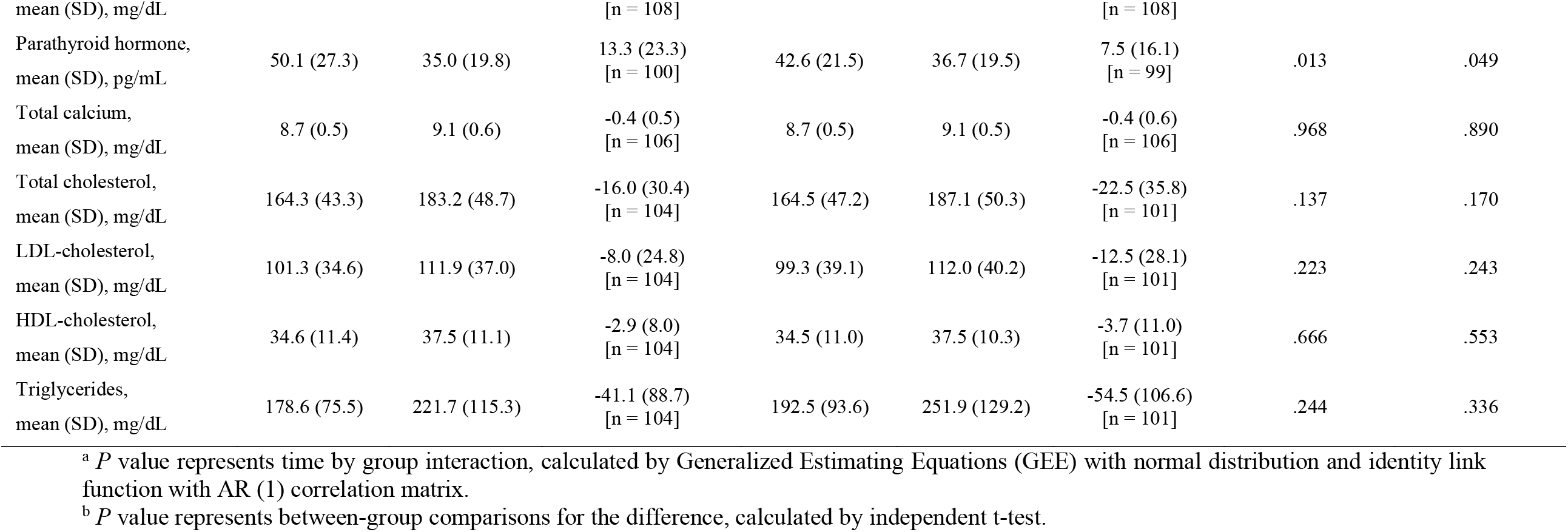
Laboratory variables.

## Discussion

This is the first randomized, double-blind, placebo-controlled trial to show that vitamin D_3_ supplementation is safe and increases 25-hydroxyvitamin D levels, but is ineffective to improve hospital length of stay or any other clinical outcomes among hospitalized patients with severe COVID-19.

Vitamin D has been postulated to play an important role on immune system, acting as a regulator of both innate and adaptative responses.^6, 19^ Observational studies have shown that 25-hydroxyvitamin D levels are associated with better clinical outcomes in respiratory diseases.^20^ Positive associations between low 25-hydroxyvitamin D levels and poor prognosis among patients with COVID-19 have also been observed.^21^ Furthermore, a small-scale, non-randomized trial demonstrated that the administration of regular boluses of vitamin D_3_ before the infection was associated with better survival and less severe disease among older, frail patients with COVID-19.^22^ In the current trial, however, a single dose of 200,000 IU of vitamin D_3_ supplementation failed to promote any clinically relevant effects among hospitalized patients with severe COVID-19, contesting the utility of supplementary vitamin D_3_ as a treatment in this disease. The lack of clinical benefits seen in this study was independent of the ability of vitamin D_3_ supplementation to increase serum 25-hydroxyvitamin D levels. In fact, following the intervention, 86.7% of the patients in the supplementation arm achieved vitamin D sufficiency (≥ 30 ng/mL) vs 11% only in the placebo group. In a sensitivity analysis confined to the patients exhibiting 25-hydroxyvitamin D deficiency, vitamin D_3_ supplementation remained effective in increasing 25-hydroxyvitamin D levels vs placebo; yet, no clinical improvements were noted. Collectively, these analyses indicate that a single oral dose of 200,000 IU of supplementation can rapidly increase 25-hydroxyvitamin levels, in agreement with our hypothesis, so that the present null findings cannot be attributed to the failure of increasing serum 25-hydroxyvitamin D levels.

Despite the clinical inefficacy of vitamin D_3_ supplementation, the intervention was not associated with any important adverse events or meaningful changes in laboratory parameters, suggesting that a relatively high-dose of vitamin D_3_ can be well tolerated in general and free of adverse effects in patients with COVID-19.

The strengths of this study include the randomized, double-blind, placebo-controlled experimental design, the adequate power, particularly for the primary analysis, the very low attrition rate (3.3%), the concomitant assessment of 25-hydroxyvitamin D levels along with clinical outcomes, and the assessment of hospitalized patients with severe COVID-19.

## Limitations

This trial has several limitations. First, the sample size could have been underpowered to detect significant changes for the secondary outcomes. Second, as the patients had several coexisting diseases and were subjected to a diverse medication regimen, the results could have been affected by the heterogeneity of the sample and its treatment. Third, the proportion of patients with 25-hydroxyvitamin D deficiency enrolled in this study was considerably lower than those reported in other cohorts,^23^ possibly as a consequence of differences in geographic locations. Although we conduced sensitivity analyses involving patients with 25-hydroxyvitamin D deficiency, one could argue that they could have been underpowered, as previously pointed out. Therefore, caution should be exercised in generalizing these findings to patients from other geographical regions. Finally, the findings should be also confined to the dose and supplementation strategy used in this trial. Further studies should determine whether preventive or early vitamin D_3_ supplementation could be useful in the treatment of patients with COVID-19, especially those with a mild or moderate disease.

## Conclusions

Among hospitalized patients with severe COVID-19, a single dose of 200,000 IU of vitamin D_3_ supplementation was safe and increased 25-hydroxyvitamin D levels, but did not reduce hospital length of stay or any other clinically relevant outcomes vs placebo. Thus, this trial does not support the use of vitamin D_3_ supplementation as an adjuvant treatment of patients with COVID-19.

## Data Availability

Data Availability Statement
Data
Data available: Yes.
Data types: Deidentified participant data.
How to access data: Requests must be sent to
When available: Four months after publication.
Supporting documents
Documents types: None.
Additional information
Who can access the data: Qualified clinical researchers.
Types of analysis: Specified purposes dependent on the nature of the request and the intention use of the data.
Mechanisms of data availability: Signed data access agreement.
Any additional restrictions: The request must include a statistician.

## Article information

### Author Contributions

Dr. Pereira had full access to all of the data in the study and take responsibility for the integrity of the data and the accuracy of the data analysis.

*Concept and design*: Murai, Fernandes, Pinto, Goessler, Gualano, Pereira.

*Acquisition, analysis and interpretation*: All authors.

*Drafting of the manuscript*: Murai, Fernandes, Gualano, Pereira.

*Critical revision of the manuscript for important intellectual content*: All authors.

*Statistical analysis*: Murai, Fernandes, Pinto, Reis, Gualano, Pereira.

*Obtained funding*: Gualano, Pereira.

*Supervision*: Gualano, Pereira.

*Administrative, technical, or material support*: Sales, Antonangelo, Caparbo.

### Data Sharing Statement

See Supplement 3.

### Additional Contributions

The authors are thankful to Dr. Monica Pinheiro and Dr. Roberta Costa for the assistance at Ibirapuera Field Hospital; Dr. Rogério Ruscitto do Prado for conducting statistical analyses; Mayara Diniz Santos for the technical support; all the staff members from both centers; all the patients who participated in this study.

---

**Supplementary Table 1.**
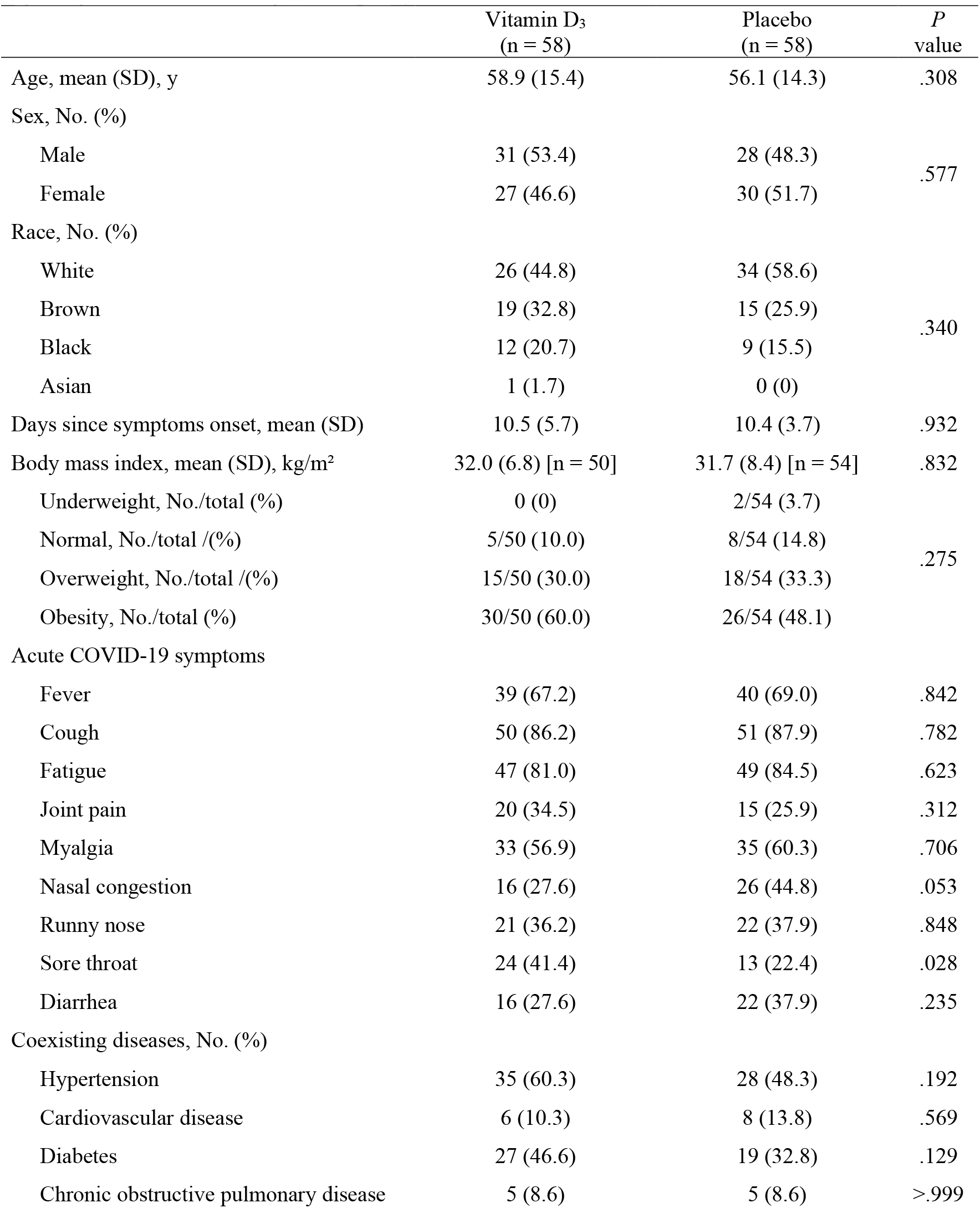
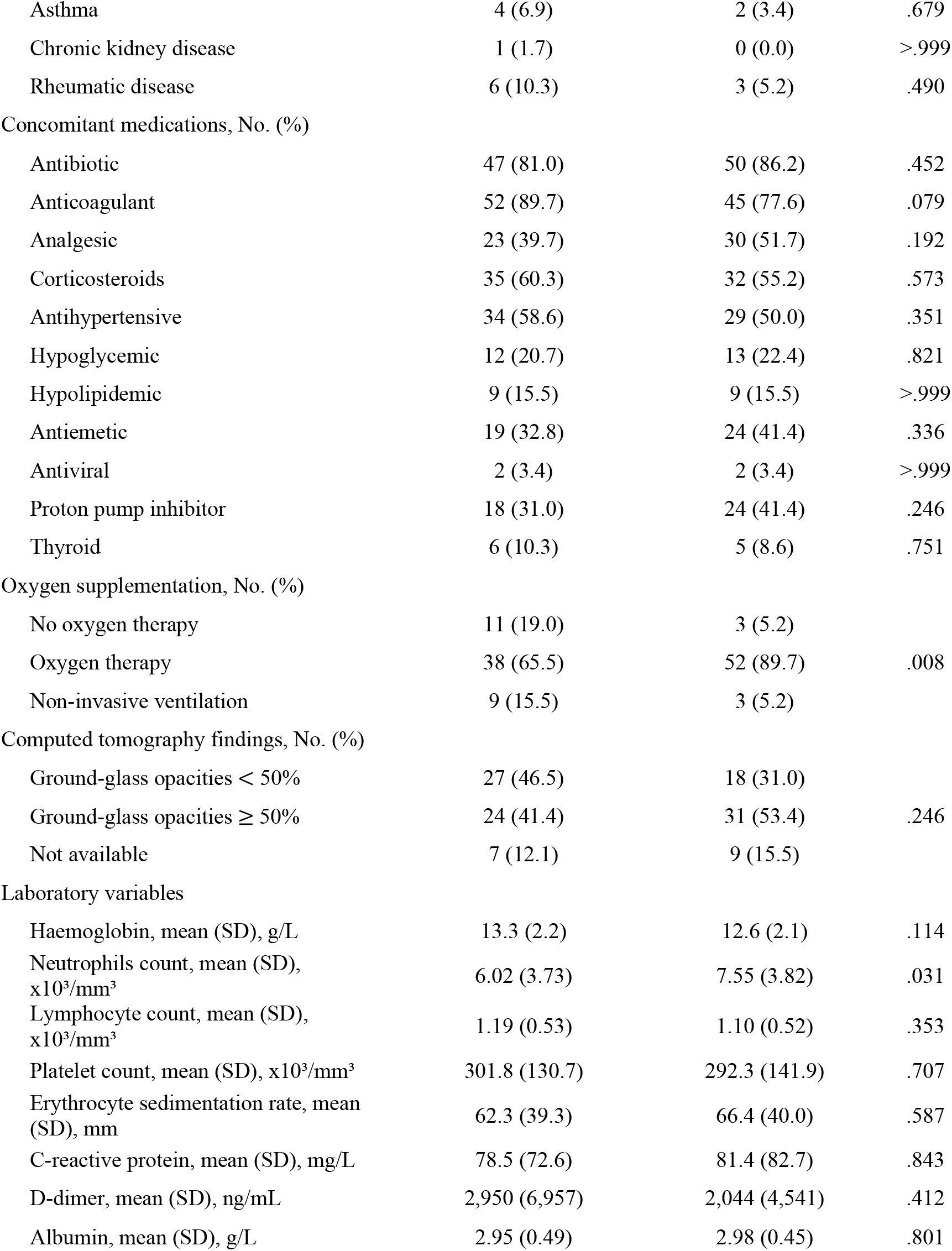
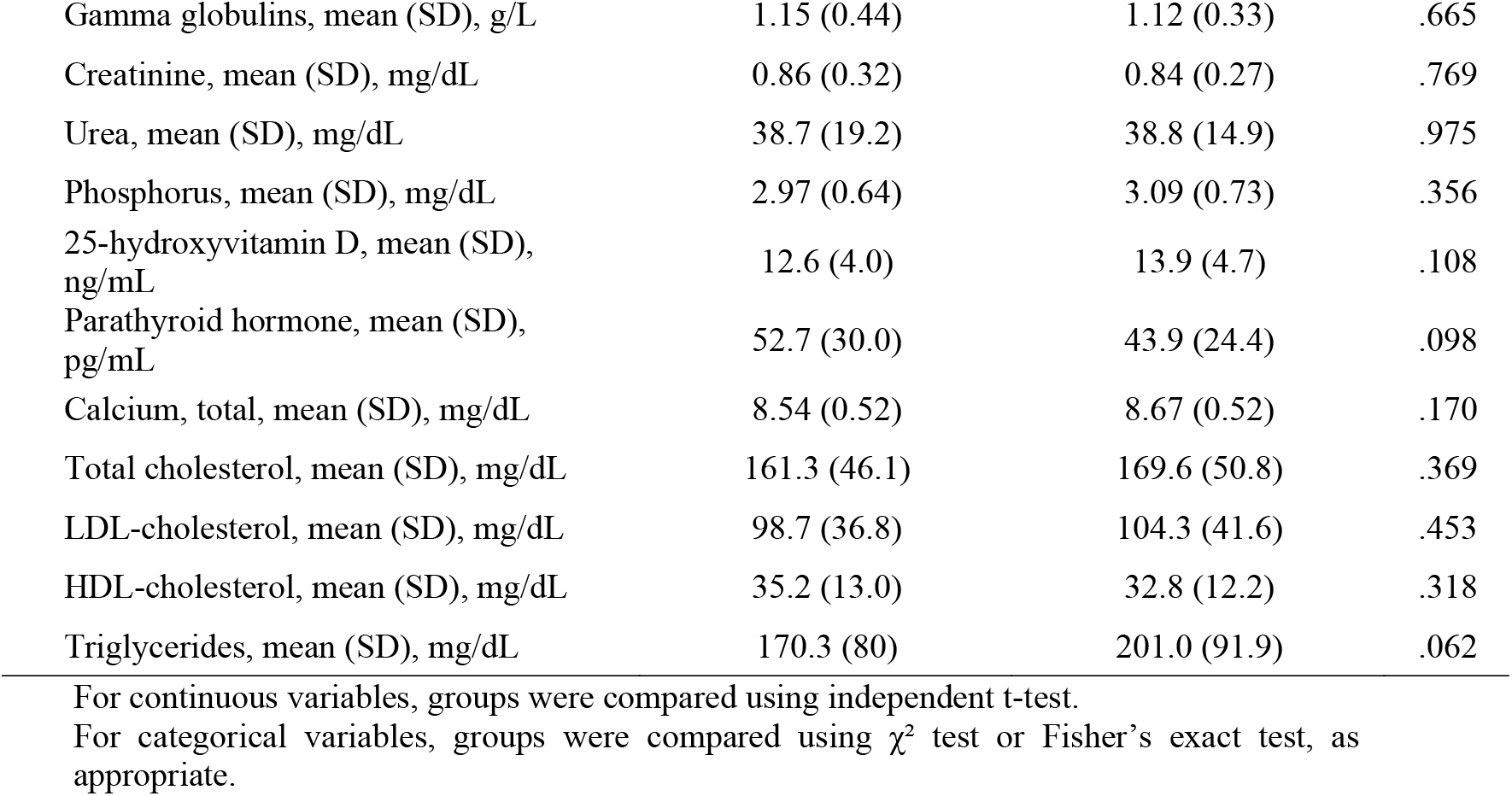
Baseline demographic and clinical characteristics from patients with 25-hydroxyvitamin D deficiency (< 20 ng/mL).

|  | Vitamin D<sub>3</sub><br>(n = 58) | Placebo<br>(n = 58) | *P*<br>value |
| --- | --- | --- | --- |
| Age, mean (SD), y | 58.9 (15.4) | 56.1 (14.3) | .308 |
| Sex, No. (%) |  |  |  |
| Male | 31 (53.4) | 28 (48.3) | .577 |
| Female | 27 (46.6) | 30 (51.7) |  |
| Race, No. (%) |  |  |  |
| White | 26 (44.8) | 34 (58.6) | .340 |
| Brown | 19 (32.8) | 15 (25.9) |  |
| Black | 12 (20.7) | 9 (15.5) |  |
| Asian | 1 (1.7) | 0 (0) |  |
| Days since symptoms onset, mean (SD) | 10.5 (5.7) | 10.4 (3.7) | .932 |
| Body mass index, mean (SD), kg/m<sup>2</sup> | 32.0 (6.8) [n = 50] | 31.7 (8.4) [n = 54] | .832 |
| Underweight, No./total (%) | 0 (0) | 2/54 (3.7) | .275 |
| Normal, No./total /(%) | 5/50 (10.0) | 8/54 (14.8) |  |
| Overweight, No./total /(%) | 15/50 (30.0) | 18/54 (33.3) |  |
| Obesity, No./total (%) | 30/50 (60.0) | 26/54 (48.1) |  |
| Acute COVID-19 symptoms |  |  |  |
| Fever | 39 (67.2) | 40 (69.0) | .842 |
| Cough | 50 (86.2) | 51 (87.9) | .782 |
| Fatigue | 47 (81.0) | 49 (84.5) | .623 |
| Joint pain | 20 (34.5) | 15 (25.9) | .312 |
| Myalgia | 33 (56.9) | 35 (60.3) | .706 |
| Nasal congestion | 16 (27.6) | 26 (44.8) | .053 |
| Runny nose | 21 (36.2) | 22 (37.9) | .848 |
| Sore throat | 24 (41.4) | 13 (22.4) | .028 |
| Diarrhea | 16 (27.6) | 22 (37.9) | .235 |
| Coexisting diseases, No. (%) |  |  |  |
| Hypertension | 35 (60.3) | 28 (48.3) | .192 |
| Cardiovascular disease | 6 (10.3) | 8 (13.8) | .569 |
| Diabetes | 27 (46.6) | 19 (32.8) | .129 |
| Chronic obstructive pulmonary disease | 5 (8.6) | 5 (8.6) | >.999 |
| Asthma | 4 (6.9) | 2 (3.4) | .679 |
| Chronic kidney disease | 1 (1.7) | 0 (0.0) | >.999 |
| Rheumatic disease | 6 (10.3) | 3 (5.2) | .490 |
| Concomitant medications, No. (%) |  |  |  |
| Antibiotic | 47 (81.0) | 50 (86.2) | .452 |
| Anticoagulant | 52 (89.7) | 45 (77.6) | .079 |
| Analgesic | 23 (39.7) | 30 (51.7) | .192 |
| Corticosteroids | 35 (60.3) | 32 (55.2) | .573 |
| Antihypertensive | 34 (58.6) | 29 (50.0) | .351 |
| Hypoglycemic | 12 (20.7) | 13 (22.4) | .821 |
| Hypolipidemic | 9 (15.5) | 9 (15.5) | >.999 |
| Antiemetic | 19 (32.8) | 24 (41.4) | .336 |
| Antiviral | 2 (3.4) | 2 (3.4) | >.999 |
| Proton pump inhibitor | 18 (31.0) | 24 (41.4) | .246 |
| Thyroid | 6 (10.3) | 5 (8.6) | .751 |
| Oxygen supplementation, No. (%) |  |  |  |
| No oxygen therapy | 11 (19.0) | 3 (5.2) |  |
| Oxygen therapy | 38 (65.5) | 52 (89.7) | .008 |
| Non-invasive ventilation | 9 (15.5) | 3 (5.2) |  |
| Computed tomography findings, No. (%) |  |  |  |
| Ground-glass opacities < 50% | 27 (46.5) | 18 (31.0) |  |
| Ground-glass opacities ≥ 50% | 24 (41.4) | 31 (53.4) | .246 |
| Not available | 7 (12.1) | 9 (15.5) |  |
| Laboratory variables |  |  |  |
| Haemoglobin, mean (SD), g/L | 13.3 (2.2) | 12.6 (2.1) | .114 |
| Neutrophils count, mean (SD),<br>x10<sup>3</sup>/mm<sup>3</sup> | 6.02 (3.73) | 7.55 (3.82) | .031 |
| Lymphocyte count, mean (SD),<br>x10<sup>3</sup>/mm<sup>3</sup> | 1.19 (0.53) | 1.10 (0.52) | .353 |
| Platelet count, mean (SD), x10<sup>3</sup>/mm<sup>3</sup> | 301.8 (130.7) | 292.3 (141.9) | .707 |
| Erythrocyte sedimentation rate, mean<br>(SD), mm | 62.3 (39.3) | 66.4 (40.0) | .587 |
| C-reactive protein, mean (SD), mg/L | 78.5 (72.6) | 81.4 (82.7) | .843 |
| D-dimer, mean (SD), ng/mL | 2,950 (6,957) | 2,044 (4,541) | .412 |
| Albumin, mean (SD), g/L | 2.95 (0.49) | 2.98 (0.45) | .801 |
| Gamma globulins, mean (SD), g/L | 1.15 (0.44) | 1.12 (0.33) | .665 |
| Creatinine, mean (SD), mg/dL | 0.86 (0.32) | 0.84 (0.27) | .769 |
| Urea, mean (SD), mg/dL | 38.7 (19.2) | 38.8 (14.9) | .975 |
| Phosphorus, mean (SD), mg/dL | 2.97 (0.64) | 3.09 (0.73) | .356 |
| 25-hydroxyvitamin D, mean (SD),<br>ng/mL | 12.6 (4.0) | 13.9 (4.7) | .108 |
| Parathyroid hormone, mean (SD),<br>pg/mL | 52.7 (30.0) | 43.9 (24.4) | .098 |
| Calcium, total, mean (SD), mg/dL | 8.54 (0.52) | 8.67 (0.52) | .170 |
| Total cholesterol, mean (SD), mg/dL | 161.3 (46.1) | 169.6 (50.8) | .369 |
| LDL-cholesterol, mean (SD), mg/dL | 98.7 (36.8) | 104.3 (41.6) | .453 |
| HDL-cholesterol, mean (SD), mg/dL | 35.2 (13.0) | 32.8 (12.2) | .318 |
| Triglycerides, mean (SD), mg/dL | 170.3 (80) | 201.0 (91.9) | .062 |

For continuous variables, groups were compared using independent t-test.
For categorical variables, groups were compared using $\chi^2$ test or Fisher's exact test, as appropriate.

